# Optimizing the COVID-19 Intervention Policy in Scotland and the Case for Testing and Tracing

**DOI:** 10.1101/2020.06.11.20128173

**Authors:** Andreas Grothey, Ken Mckinnon

**Affiliations:** School of Mathematics, University of Edinburgh, UK

## Abstract

Unlike other European countries the UK has abandoned widespread testing and tracing of known SARS-CoV-2 carriers in mid-March. The reason given was that the pandemic was out of control and with wide community based spread it would not be possible to contain it by tracing any longer. Like other countries the UK has since relied on a lockdown as the main measure to contain the virus (or more precisely the reproduction number *ℛ*) at significant economic and social cost. It is clear that this level of lockdown cannot be sustained until a vaccine is available, yet it is not clear what an exit strategy would look like that avoids the danger of a second (or subsequent waves).

In this paper we argue that, when used within a portfolio of intervention strategies, widespread testing and tracing leads to significant cost savings compared to using lockdown measures alone. While the effect is most pronounced if a large proportion of the infectious population can be identified and their contacts traced, under reasonable assumptions there are still significant savings even if the fraction of infectious people found by tracing is small.

We also present a policy optimization model that finds, for given assumptions on the disease parameters, the best intervention strategy to contain the virus by varying the degree of tracing and lockdown measure (and vaccination once that option is available) over time. We run the model on data fitted to the published COVID-19 outbreak figures for Scotland. The model suggests an intervention strategy that keeps the number of COVID-19 deaths low using a combination of tracing and lockdown. This strategy would only require lockdown measures equivalent to a reduction of *ℛ* to about 1.8–2.0 if lockdown was used alone, at acceptable economic cost, while the model finds no such strategy without tracing enabled.

## Introduction

Unlike other European countries Scotland, like the rest of the UK, has abandoned widespread testing and tracing of known SARS-CoV-2 carriers in mid-March. The reason given was that the pandemic was out of control and with wide community based spread it would not be possible to contain it by tracing any longer. This was in line with the UK response moving from the “contain” to the “delay” phase [2]. Recently there has been talk about using a tracing app and if this could be used to contain the spread of the virus (or at least help to lower the *ℛ* number). So far such an app has not seen use in any European country, although it is an integral part of the fight against the virus in a number of Asian countries. There has also been academic study of the impact of contact tracing to the transmission of the virus in the UK [3].

This report sets out the mathematical argument for using testing and tracing, showing that for plausible assumptions about the dynamics of the virus spread and costs, when used in combination with lockdown measures, tracing leads to significant costs savings compared to using lockdown measures alone. While the effect is more pronounced if a large proportion of the infectious population can be identified and their contacts traced and isolated, it is still worthwhile even if the fraction is smaller (for example since many people may be asymptomatic).

We also present a policy optimization model that finds the best intervention strategy (as a combination of testing and tracing, lockdown and vaccination when available) dynamically over time in order to contain the spread of the virus. We use this model, calibrated to the published data on the disease spread in Scotland, to analyse the resulting economic and societal costs under different assumptions regarding limits on final death toll and tracing effectiveness. As far as we are away this is the first model presented in the literature that is optimizing the intervention strategy (rather than simulating different strategies). We find that even if tracing is limited by low effectiveness and low number of tracers, used in combination with reduced lockdown, it is still far more cost-effective than lockdown measures alone, and, if aiming to limit the final death toll, the model consistently finds better solutions in terms of death toll and costs using tracing than without it.

In the next section we give a simple mathematical model of the effect of tracing and show that, for reasonable assumptions on tracing costs and effectiveness, tracing is far more effective at reducing the ℛ number than other measure such as lockdown. In the remainder of the report we use tracing within an extended SEIR model of disease spread that is aimed at finding the optimal intervention strategy in order to contain the epidemic. In Section 3 we present the SEIR model, our extensions and the parameter fitting to the available Scottish data. In Section 4 we present the policy optimization model based on the SEIR model, while in Section 5 we present the optimal policies obtained from the model under various assumptions regarding the trade-off between economic damage and lives lost as well as limits on the acceptable death rates. In Section 6 we draw our conclusions.

## 2. The Case For Testing And Tracing

Below we show the mathematical form of a simple testing and tracing strategy for depressing the reproduction number ℛ. By testing and tracing we refer to a protocol in which some or all of the suspected (symptomatic) carriers of the virus are tested and, if positive, their past contacts over their likely infectious period are identified, tested as well, and, if necessary isolated. It is assumed that by adopting this strategy most identified carriers of the disease can be effectively eliminated as far as the disease dynamics are concerned, at least as long as the tracing is done fast enough or continued iteratively[10]. There are obvious limits to the success of this strategy in terms of being able to find a large enough number of infectious people, having the resources (in terms of test capacity and manpower) to test them and all their contacts and being able to do so in a timely manner. It is also clear that typically contact tracing will not be 100% effective: some potential infectious contacts are likely to be missed. As far as this report is concerned we do not distinguish if tracing is done by an app or ‘by hand’, although there is evidence that tracing by app has clear advantages since the response is faster [3].

We first illustrate the approach with a concrete scenario: assume that there are *I* = 2500 people currently infectious out of a population of *N* (this would correspond roughly to the number of infectious people in Scotland on 11 May according to our model fit presented in Section 3). Further assume that we know (by testing) 1000 of them and trace their contacts. This means we identify anyone that has been in contact with any of these 1000 during their period of infectiousness and isolate them (at least until they themselves can be tested and confirmed to not carry the virus), preventing them from spreading the virus further. Tracing will not be 100% effective but we assume that it can reduce the amount of spreading for a tested and traced person by 90%: *i*.*e*. rather than infecting ℛ new people, if contacts are traced this person will only infect ℛ/10 people. We assume that each infectious person will have on average 25 contacts that need to be traced (Kucharski et al[11] report 20–30 contacts on average). Assuming ℛ = 2.5, if testing and tracing is done for 1000 out of 2500 infectious people, the 2500 would infect over one generation of the virus another

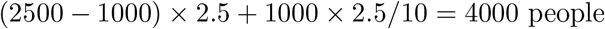

this corresponds to an effective ℛ number of ℛ^*TR*^ = 4000*/*2500 = 1.6 or a reduction in ℛ to a fraction *σ*^*TR*^ = 0.64 of its previous value.

In general assume that out of a population of *N*, there are *I* people infectious with a reproduction number of ℛ. Assume that *T* of the infectious people are identified and their contacts traced and isolate and that this reduces the reproduction rate for those *T* people to *η ℛ, i*.*e*.. the effectiveness of tracing is (1 *− η*). In that case the *I* infectious people will infect another (*I − T*)ℛ + *ηT*ℛ after one generation, compared to *I*ℛ without tracing, corresponding to reduction in infection rate of

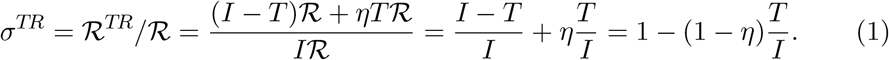

Note that the right-hand side is independent of ℛ and *N*. It is non-obvious what cost should be assigned to tracing. On the one hand we can assume that an identified contact is tested and only if positive told to isolate. If we assume a cost of £50 per test, that would give a total cost of *cost*_*trace*_ = 25 *× £*50 = *£*1250 to trace and test all contacts of an identified infectious person with cost for the tracer and isolation being neglected. This is likely to an underestimate of the true cost. On the other hand we could assume that contacts are not tested but told to isolate for 14 days (essentially enforcing a lockdown only for these people). The lockdown of the total population in the UK has reportedly led to a reduction of GDP for the effected time of 30% (£2.4bn per day out of a GDP per day of £7.9 in the UK [6]). Scotland has a GDP of £85 per person and day. If we assume the loss of 30% of the GDP contribution of the 25 contacts over 14 days we arrive at a cost of *cost*_*trace*_ = 0.3 *×* 25 *×* 14 *× £*85 = *£*8950. For the remainder of this report we assume an intermediate cost of *cost*_*trace*_ = *£*6500 per initial infection identified, to test, find all contacts and isolate them as long as necessary. Using this value, testing and contact tracing for the 1000 known infectious people would amount to a cost of 1000 *× £*6500 = *£*6.5 million.

We try to work out the cost of achieving a similar reduction *σ*^*LD*^ in ℛ by lockdown measures. The current lockdown in the UK seems to have achieved a reduction of ℛ to about 0.85 from 2.5 (which equals to a relative reduction of *σ*^*LD*^ = 0.34) and has led to a reduction of GDP for the effected time of 30%[6]. If we assume a linear relationship between lockdown level and impact on the economy (also assuming that *σ*^*LD*^ = 1, corresponding to no lockdown, also results in no economic damage) we would get the (daily) cost of lockdown as

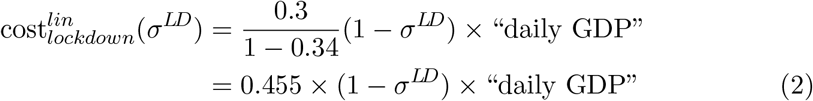

This is likely overly pessimistic for small reductions in ℛ, since there are easy measures (hand-washing, wearing of masks, mild social distancing) that can be adopted with minimal impact on the economy. In general the higher the required impact on ℛ, the more costly measures have to be taken. This leads us to assume a convex cost function, in particular we suggest to fit a quadratic. If we use the fixed points of cost_*lockdown*_(1) = 0 and cost_*lockdown*_(0.34) = 0.3 together with a flat tangent at *σ* = 1 we arrive at the daily cost function

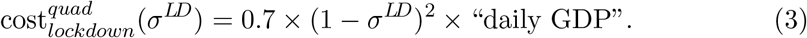

In fact the coefficient should be 0.689 which we have rounded to 0.7.

Scotland has a daily GDP of £450 million. A lockdown level corresponding to the reduction achieved by tracing *σ*^*LD*^ = *σ*^*TR*^ = 0.64 over one generation of the virus (*t*^*Gen*^ = 4.5 days [8]) would therefore result in a loss to the economy of £184 million which is almost 30 times higher than the cost of tracing.

There are obviously quite a few assumptions in this calculation and we can ask how robust these results are if we change the assumptions. First note that the analysis given does not depend on the underlying reproduction number ℛ. As ℛ does not appear on the right side of (1), if we had assumed a different value it would lead to the same *σ*^*TR*^ and subsequently the same cost of tracing and cost of achieving the equivalent reduction in ℛ by lockdown.

On the other hand, if there were 25, 000 (rather than 2500) infectious people of which we can trace 1000 then the number of new infections over one generation would amount to

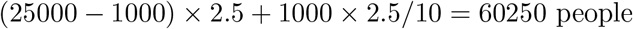

corresponding to an ℛ^*TR*^ = 2.41 or a reduction in ℛ by a factor *σ*^*TR*^ = 0.964. According to (3) the economic cost of achieving this reduction by lockdown over one generation of the virus would only be £1.8 million which is now cheaper than the cost of tracing. While tracing is much cheaper than lockdown if a reasonably large number of the infectious can be traced, there is a tradeoff and at some point the cost of lockdown will become cheaper than the cost of tracing if a too small proportion of the infectious are traced.

In practice tracing will, however, not be used in isolation but as one part of an intervention portfolio to contain the virus, with other factors suppressing the ℛ number such as lockdown measures and the build-up of herd immunity. If we assume a reduction in ℛ due to lockdown measure of *σ*^*LD*^ then the effective observed reproduction number of the disease is

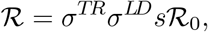

where the factor *s* is the proportion of the population that is (still) susceptible to the virus and represents the build-up of herd immunity. If the aim of the combined intervention strategy is to reduce the effective reproduction number to ℛ 1, then any reduction that is not achieved by tracing would need to be achieved by lockdown:

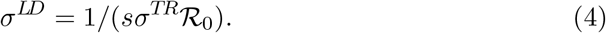

Using the *σ*^*TR*^ = *σ*^*TR*^(*T, η, I*) from (1) in (4) and the costs of tracing (*cost*_*trace*_ *× T*) and lockdown (3) we can deduce the total cost of reducing ℛ from ℛ_0_ to 1 by a combination of tracing and lockdown if *T* out of *I* infectious people are traced with an effectiveness *η* over one generation of the virus to be

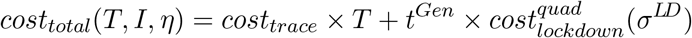

Below we plot this cost curve as a function of the proportion of infectious traced (*T/I*) for different assumed numbers of infectious people *I* in the population. The plot on the left assumes no herd immunity (*s* = 1), whereas in the plot on the right 20% of the population are immune (*s* = 0.8). In these plots no tracing would correspond the the values on the right edge (*T/I* = 0).

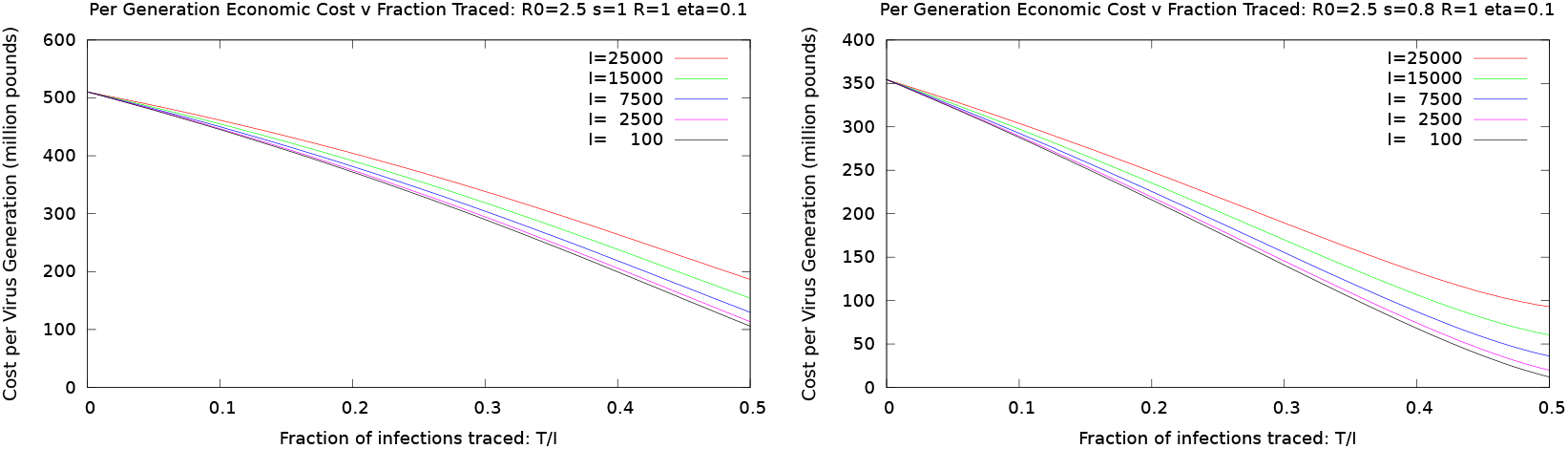

We see that in the combined policy there is a clear advantage of tracing as part of a combined portfolio almost irrespective of the number of infectious people in the population. The cost advantage becomes more pronounced the higher the proportion of traced people is, decreasing the total cost by about a factor of 4 −− 5 for *T/I* = 0.5 in the *s* = 1 case and 7 −− 10 in the *s* = 0.8 case, however it is present for any assumed proportion. We should note that there are obvious simplification in this static, over one virus generation, analysis of the effect and cost of tracing. In the remainder of the report we will look at tracing as part of a portfolio of intervention strategies that is optimized dynamically for every point in time.

Since it will be useful to interpret the later results we give a graph of the reduction in lockdown *σ*^*LD*^ (expressed as ℛ^*LD*^ = *σ*^*LD*^ ℛ_0_, the effective ℛ if lockdown were used on its own) needed to contain the virus as a function of the tracing parameters *T/I* and *η*, according to (4) and (1).

In the bottom right corner (below the ℛ^*LD*^ = 2.5 contour), tracing alone is enough to contain the virus, anywhere else the indicated ℛ would need to be achieved by other measures such as lockdown.

In the remainder of this report we will explore the effectiveness of using contact tracing as part of a policy optimization model that controls ℛ in order to limit the spread of the disease.

## 3 The Extended SEIR Model And Data Fitting

We are using an extension of the SEIR model[1]. The SEIR model is a compartmental dynamic model of disease spread, in which *S*_*t*_ is the number of susceptible people at time *t, E*_*t*_ is the exposed (*i*.*e*. infected with the virus), *I*_*t*_ are the people currently infectious and *R*_*t*_ is the recovered (*i*.*e*. no longer infectious. ‘Recovered’ does include people that have died. It does not necessarily mean that people have completely recovered from the illness, only that they are no longer spreading the virus. The SEIR model uses infection dynamics according to the difference equations

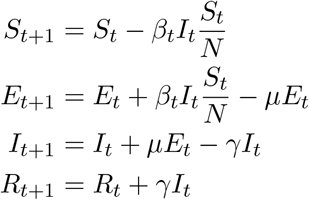

Here *β*_*t*_, *µ* and *γ* are the (daily) rates of infection, incubation, and recovery. That is the time of an individual moving from exposed to infectious state is assumed to be exponentially distributed with a mean incubation period of *t*^*µ*^ = 1*/µ* days. For the transition between *S* and *E* it is assumed that every infectious person *I*_*t*_ infects on average *β*_*t*_ new people per day if there is no immunity in the population, multiplied by the fraction *S*_*t*_*/N* if some people have already been removed from the pool *S*_*t*_ of infectious people. Since an individual remains infectious for 1*/γ* days, they will infect on average ℛ_0_ = *β/γ* individuals. ℛ_0_ is the basic reproduction number of the disease.^1^ When all the population is susceptible the number of infected individuals will increase by a factor of ℛ_0_ with every generation of the virus. The generation time *t*^*Gen*^ is the average time from infection to passing the virus on. We assume that *β*_*t*_ can vary with time *t*, whereas *γ* and *µ* are constant. This model assumes that people that have recovered from the disease are immune to it and that the immunity will last for the length of the modelling period.

The SEIR model does not keep track of the fate of the infected individuals: namely if they require hospital or intensive care (ICU) treatment, nor in fact whether the individuals will ultimately die or recover. In order to take these into account we extend the SEIR model as follows:

- Any exposed individual has a chance of *ϕ*_*h*_ of needing hospital treatment. Of those exposed individuals that will ultimately go to hospital a fraction *r*_*h*_ will do so per day (that is they take an average of *t*^*hr*^ = 1*/r*_*h*_ days from becoming exposed to needing hospital treatment).
- Any individual in hospital has a chance of *ϕ*_*icu*_ of needing to go into ICU. If so, a fraction *r*_*icu*_ will do so per day (that is they take an average of *t*^*icur*^ = 1*/r*_*icu*_ days from being admitted to hospital to needing intensive care). Of those not going to ICU eventually, an average fraction of *r*_*h*_ will recover each day (that is they spend on average *t*^*h*^ = 1*/r*_*h*_ days in hospital).
- Any individual in ICU has a chance of *ϕ*_*d*_ of dying. Of those that do, a fraction *r*_*d*_ will die per day (that is they take an average of *t*^*d*^ = 1*/r*_*d*_ days from being admitted to ICU until dying). Of those not dying eventually, an average fraction of *r*_*icu*_ will recover per day (that is they spend on average *t*^*icu*^ = 1*/r*_*icu*_ days in ICU).
- There is limited hospital and ICU capacity. Individuals that would need hospital or ICU when there is no more capacity will die.
- The model makes the following simplifications: apart from the above hospital and ICU rejects, nobody will die that has not gone through the hospital and ICU route. The model does not track community or care home deaths. People recovered from ICU will not go back to hospital but leave the model.
- The model does not discriminate by age groups.
- There is no import of cases (for example from other countries).

There is a relation of the above parameters to the infection fatality rate, (*i*.*e*. the fraction of those exposed to the virus that will ultimately die) by

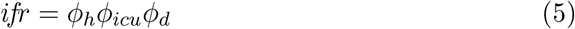

In order to model the effect of lockdown (and other measures to control the infection rate) we use a time dependent ℛ rate ℛ_*t*_. We assume that initially the ℛ rate was ℛ_*t*_ = ℛ_0_ = 2.5 (as reported in various studies) and after the initial lockdown decreased to a constant value 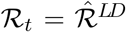. This time-varying ℛ_*t*_ affects the dynamics of the disease evolution through *β*_*t*_ = ℛ_*t*_*γ*.

### 3.1. Parameter Fitting

We have used the model as above for a nonlinear least squares parameter fit to the available COVID-19 data for Scotland. We have used data published by the Scottish Government for the period from 18 March to 11 May 2020[9]. The data available is number of positive tests, number of hospital admissions (separated by tested and suspected), number of patients in intensive care (again tested and suspected) and number of deaths. We have fitted our extended SEIR model to the numbers of hospital admissions (total of tested and suspected), ICU admissions (again total of tested and suspected) and deaths. We have decided not to use the number of positive tests since a) the access to tests to various sections of society has changed over time repeatedly and b) it is widely believed that there is a large amount of undertesting (i.e. a large and unknown fraction of infected people are not tested and therefore not detected).

The unknown rate of undertesting poses problems for the model fitting. There are conceivably many different fits to the available data corresponding to a different proportion of the population that has already been exposed to the disease[12], ultimately leading to different estimates of people needing hospital treatment *ϕ*_*h*_ and a different infection fatality rate *ifr*. We are dealing with this problem by setting an *ifr* = 0.6% and using this to constrain parameters *ϕ*_*h*_, *ϕ*_*icu*_ and *ϕ*_*d*_ via (5). The study in [15] from mainland China arrives at at estimate of *ifr* = 0.6%. Estimates of different studies on the *ifr* by different groups seem to converge on an interval of *ifr* ∈ [0.3%, 1%]. Also using *ifr* = 0.6% in our model results in a fit by which about 7.5% of the Scottish Population have been exposed to the virus on 11 May which seems roughly in line with the values reported by various studies on achieved immunity levels in different countries. Note that assuming *ifr* = 0.3% will result in a fit where about twice as many people have already been infected whereas for an *ifr* = 1.2% about half as many people would have been affected. Changing *ifr* has a proportional effect on the hospitalisation rate *ϕ*_*h*_ with all other model parameters staying roughly the same.

We further set a basic reproduction rate before lockdown of ℛ_0_ = 2.5 as assumed in various studies. We have tried to use the least squares fit to determine the best ℛ_0_. However, the parameter fitting model is near degenerate with respect to ℛ_0_, that is there are almost equally good fits with different values for ℛ_0_. Different rates for ℛ_0_ lead to different values *µ* and 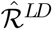 while other parameters are remarkably similar. We have chosen to go with the fit for ℛ_0_ = 2.5 which corresponds to an 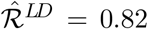 which is close to quoted estimates by the Scottish Government. In Table 1 we state the model parameters obtained by our fits.

**Table 1.**
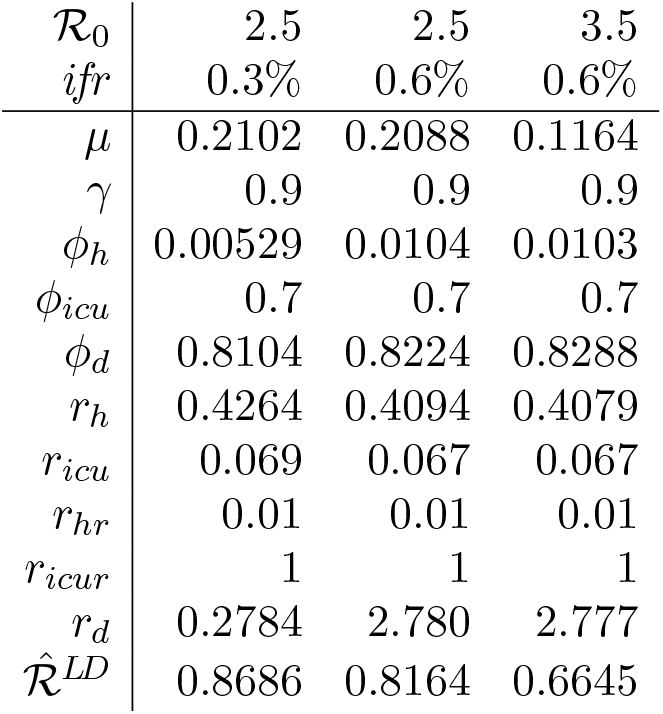
Best fits of SEIR parameters for different assumed values of *ifr* and ℛ_0_.

## 4. Policy Optimization

In this section we describe how we use our fitted SEIR model to determine the best policy for controlling the spread of the disease forward from the date of fitting the data^2^. The control options we are considering are

- Lockdown,
- Testing and tracing,
- Vaccination (assumed to be only available after day *t* = 400).

The optimization will aim to determine the optimal amount of each of these options per day, with respect to several different objectives and constraints (on amount of available tracing and acceptable number of deaths). We discuss the impact and changes to the model for these below.

### 4.1. Lockdown

For any time *t* going forward from the current time we introduce a decision variable 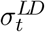 that controls the suppression of ℛ_0_ achieved by lockdown. It links the effective reproduction rate 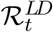 at time *t* achieved by lockdown measures (but not taking account of the build up of immunity) to the basic reproduction rate ℛ_0_ through

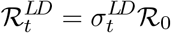

We will assume that 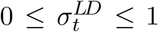 and that 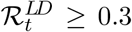 (ℛ = 0.3 was estimated to be the rate in Wuhan during the tightest lockdown, which we assume to be the maximum achievable by this measure). As the economic (daily) cost of the lockdown we use

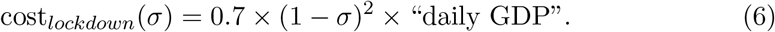

as outlined in Section 2.

### 4.2. Testing and Tracing

The Testing and Tracing protocol used in the model assumes that once a person is detected to be infectious, that person and all their (identified) contacts are isolated, thus effectively removing that person from the exposed population before they infect anyone else (in fact tracing is usually done after others are infected, but since these are also isolated the effect on the disease dynamics is as if the infectious person is removed before passing it on). Tracing will not be 100% effective, so we assume that rather than completely eliminating a person from the pool of exposed we are able to decrease their infectiousness from *β* to some fraction, say *ηβ* (with default *η* = 0.1). As outlined earlier we assume a cost of cost_*trace*_ = *£*6500 to decrease the infectiousness of one person by testing and tracing. There will be a maximum capacity of testing and tracing which is determined by a) the number of available tracers and b) the number of tests that can be done within one day. We set a default limit of 900 on the number people that can have their contacts traced in Scotland per day. Further, it is not possible to find (and eliminate) every infectious person this way (since some people may only have very mild or no symptoms). We therefore assume that we can only eliminate up to 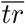 (default 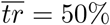) of the newly infectious people by tracing. In the model each newly infectious person is assigned to either a *traceable* or *untraceable* compartment depending on the probability indicated by 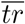.

### 4.3. Vaccination

We are aiming to use our model to find the best intervention strategy from now until a vaccine is found. We assume this time horizon to be *T*_*vac*_ = 300 days after the start of the policy optimization period^3^, which would be early March 2021). If we simply stop the policy optimization at time *T*_*vac*_, the model will attempt to relax the lockdown (and start a second wave) just before the end of the modelling period since it cannot see the deaths caused by this second wave after the modelling horizon. In order to overcome this problem we assume that a vaccination programme runs from day *t* = 301 to *t* = 400 which would be able to vaccinate 1% of the population on every one of these 100 days. We assume it will not be known which people are immune at that point and which not, so the vaccinations will be given to all people equally although people will not be vaccinated twice. In the model we allow the removal of a fraction of the susceptible population (the S compartment) corresponding to 1% of the susceptible population each day after day *t* = 300. We assume no cost is associated with this.

### 4.4. Cost of Deaths

In order to do a tradeoff between the economic costs imposed by controlling measures and the loss of life if no measures are taken we use the concept of cost per Quality-of-life Adjusted Life Year (QALY). This concept is used in various health systems to prioritise different treatment options when resources are limited. In the UK this is done for the NHS through the National Institute for Health and Care Excellence (NICE). They suggest a QALY value in the range of £20,000 – £30,000[13]. In the model we will use £30,000 initially. Table 2 gives a breakdown of Covid infection fatality rate as estimated by [15] together with a breakdown of UK population and UK life expectancy by age brackets. Using this data implies an average of 6.9 life years lost per Covid death. This is likely an overestimate since Covid seems to badly affect mainly people with underlying health conditions which would be expected to have a lower life expectancy than the average of their age bracket. Also these are 6.9 raw life years rather than quality adjusted life years (QALY). Nevertheless we will use this higher value which gives a cost per life lost of

**Table 2.** Infection fatality rate [15] and breakdown of UK population by age bracket.

| Age bracket | % of UK population | Expected future years | IFR (%) |
| --- | --- | --- | --- |
| 0–9 | 12.1 | 75 | 0.002 |
| 10–19 | 11.3 | 65 | 0.007 |
| 20–29 | 13.1 | 55 | 0.031 |
| 30–39 | 13.3 | 45 | 0.084 |
| 40–49 | 12.8 | 36 | 0.161 |
| 50–59 | 13.5 | 27 | 0.595 |
| 60–69 | 10.6 | 18 | 1.93 |
| 70–79 | 8.3 | 11 | 4.28 |
| 80+ | 4.9 | 6 | 7.8 |

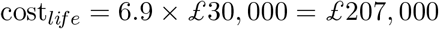

The policy optimization model is a nonlinear nonconvex constrained optimization problem. The decision variables are the severity of lockdown *σ*_*t*_, the number of people traced *T*_*t*_ (both for all times from now *t* = 1 until the time horizon *T* = 400) and the number of people vaccinated *V*_*t*_ (for *t* ≥ 301). The model keeps track of the SEIR dynamics *S*_*t*_, *E*_*t*_, *I*_*t*_, *R*_*t*_ as well as the number of people in hospital, ICU and those that have died for all time periods *t* = 1, .., *T*. We use an ICU limit of 700 beds and a total hospital capacity of 4, 200 beds which are taken from [14].

## 5. Results

We have implemented the parameter fitting and policy optimization model in AMPL[7] and solved it using FilterSQP[5] (for the parameter fitting) and IPOpt[16] (for the policy optimization). Both problems are nonlinear nonconvex constrained optimization problems. The policy optimization problem has 7894 variables and 6501 variables. IPOpt takes generally less than 1 min to solve the policy optimization model on a Linux PC with an Intel Xeon CPU running at 2.60GHz.

As a first baseline experiment we have used the policy optimization model to find the optimal intervention strategy without tracing. We use the quadratic impact to the economy according to (3), a cost per QALY of £30,000 and a vaccination programme that starts at day *t* = 301 (beginning of March 2021). The resulting optimal intervention and resulting disease dynamics are plotted in the graph below on the left

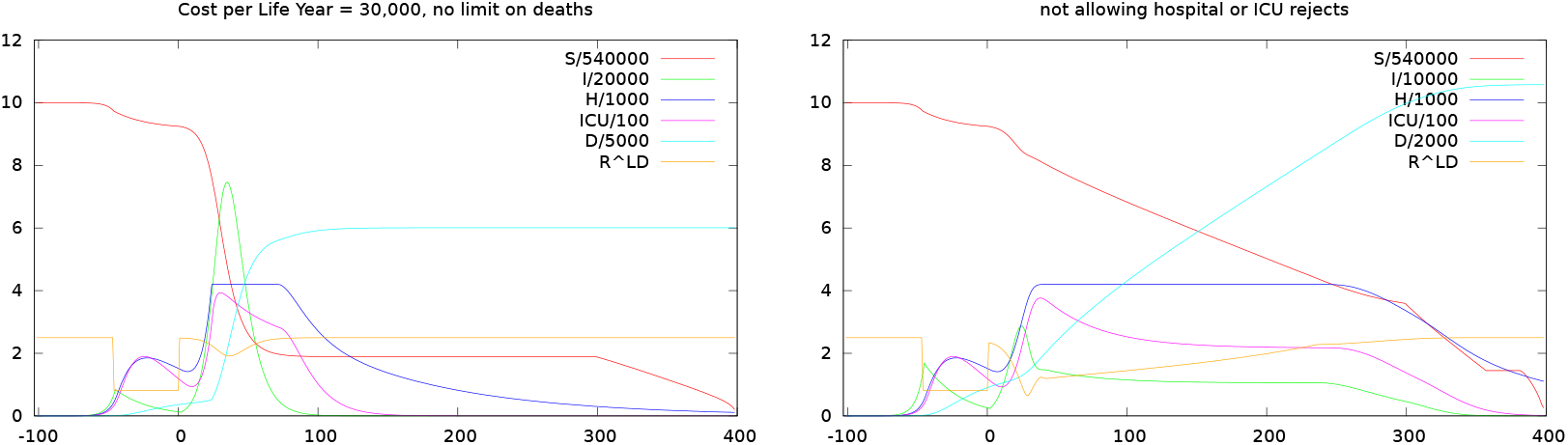

The curves plotted are the number of susceptible people (those that have not yet had contact with the virus) in the population (*S*, scaled so that 10 corresponds to the whole Scottish population), the number of currently infectious people (*I*), the number of COVID patients currently in hospital (*H*) and in ICU, and the cumulative number of deaths (*D*). In addition we plot the optimal intervention strategy, namely the amount of lockdown. The amount of lockdown is expressed as the ℛ rate that would results from this measure alone, *i*.*e*. a lockdown that suppresses *σ*^*LD*^ = 50% of infections would be plotted as ℛ^*LD*^ = *σ*^*LD*^ℛ_0_ = 0.5 × 2.5 = 1.25. The vaccination programme is not plotted separately but is in all cases used from *t* = 301 to *t* = 400 and will result in a steady removal of the population that is still susceptible at *t* = 300.

The policy optimization starts at day *t* = 1 (12 May). For *t* ≤ 0 the disease dynamics are as determined by the least squares fit and will be the same for all following diagrams. Note that at day *t* = *−* 50 (23 March) the lockdown in the UK and Scotland has started, reducing ℛ from ℛ_0_ = 2.5 to ℛ^*LD*^ = 0.81.

We see that the model decides on a herd immunity strategy as the optimal intervention: It starts by almost immediately relaxing the lockdown to 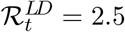. Hospitals reach capacity at day *t* = 24 and from then on until day *t* = 80 they stay at capacity and any further would be admissions are rejected and die with probability *ϕ*_*icu*_. ICU on the other hand never reach capacity. There is only a very slight re-enactment of lockdown measures around *t* = 30 to dampen the peak of the hospital rejects slightly. Nevertheless this leads to a total of 30, 322 hospital rejects and 30, 042 deaths. The total cost of this scenario if £6.586bn consisting of £6.218bn for the lives lost and £0.367bn for the economic cost of the lockdown. We see that under strictly economic objectives the model has only a very limited incentive to prevent hospitals from overflowing. Such a policy would no doubt not be politically defendable, so from now on we impose that the hospital limit of 4200 beds and the ICU limit of 700 beds is never exceeded. Re-running the model with these constraints results in the plot above on the right.

We see that the model still decides on a herd immunity strategy although this time it uses a carefully timed second lockdown: It again starts by relaxing the lockdown to 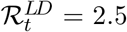 before gradually enforcing it again over the next 4 weeks, to arrive at an 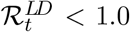 at day 24 in order to prevent the hospitals from over-flowing their capacity. It keeps a severe lockdown level of 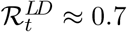 for another 10 days, before starting to release the lockdown gradually as susceptibility decreases, with 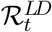 reaching 2.0 at about day 200. Ultimately the model has reached herd immunity levels before day 300 (at *t* = 300 about 65% of the population has been exposed to the virus) when the vaccination programme could be started, which is therefore not needed anymore. Note that the lockdown is only enforced in order to keep the hospital occupancy level below its capacity limit. Once that is guaranteed, the model aims to achieve herd immunity as quickly as possible in order to limit the economic cost of the lockdown. This still results in a total death toll for Scotland of around 21, 000 people. The total cost of this strategy is £15.836bn (or about 9.3% of the Scottish annual GDP), consisting of £4.379bn as the cost of the 21,000 deaths and £11.457bn as the economic impact of the lockdown.

While this is in some sense the optimal strategy given our assumptions on the relative costs of lockdown and lives lost while not exceeding hospital capacities, it is obviously problematic for a number of reasons: Firstly, even if optimal by a cost-benefit analysis the total of 21,000 deaths may be difficult to defend politically and ethically, if these deaths could be reasonably avoided. Secondly the death toll could turn out significantly higher if the assumptions in the model turn out to be wrong and hospitals are overloaded after all. This could also happen if the crucial timing of the second lockdown is not exactly right, something that would seem very difficult to achieve in practice.

To guide the model towards a solution with a lower death toll we have two possibilities: either to increase the assume value per QALY, or to limit the number of acceptable deaths.

To explore the first of these we have successively increased the assumed costs of a life year to increasing multiples of £60,000 up to a final value of £300,000. Below we give the plot of the optimal intervention strategy and the resulting disease dynamics for the final two of these.

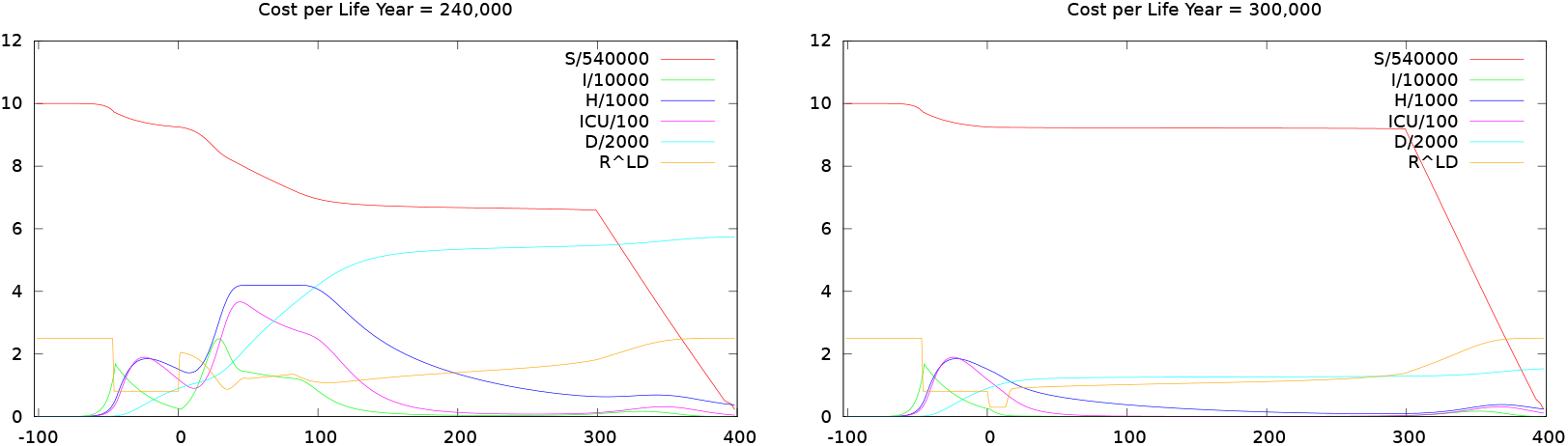

Up to £240,000 there is no significant change in the shape of the intervention 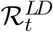. The model is still aiming to release lockdown as fast as possible, although a slightly more severe lockdown results in fewer lives lost (but still around 11, 000) and by time *t* = 300 herd immunity has not quite been achieved, so that the vaccination programme removes the remaining people from the susceptible population.

Things look differently when the cost of QALY reaches £300,000 (10 times its original level). Only then is the optimal intervention policy to keep a lockdown in place to ensure ℛ ≈ 1 until a vaccine is found. This action limits final Covid deaths to about 3, 000 but with enormous costs. The total cost (economic + life years lost) increases to £42.7bn (about 25% of the Scottish yearly GDP). Note that at this high assumed cost per QALY it is a valid question to ask whether there are other (non-Covid) interventions both within and outside the health system that would result in more life years saved with less resources.

The other option discussed above is to impose a limit on acceptable deaths. The plot below shows the results if we again use a value per QALY of £30,000 but impose a limit of deaths of 3000.

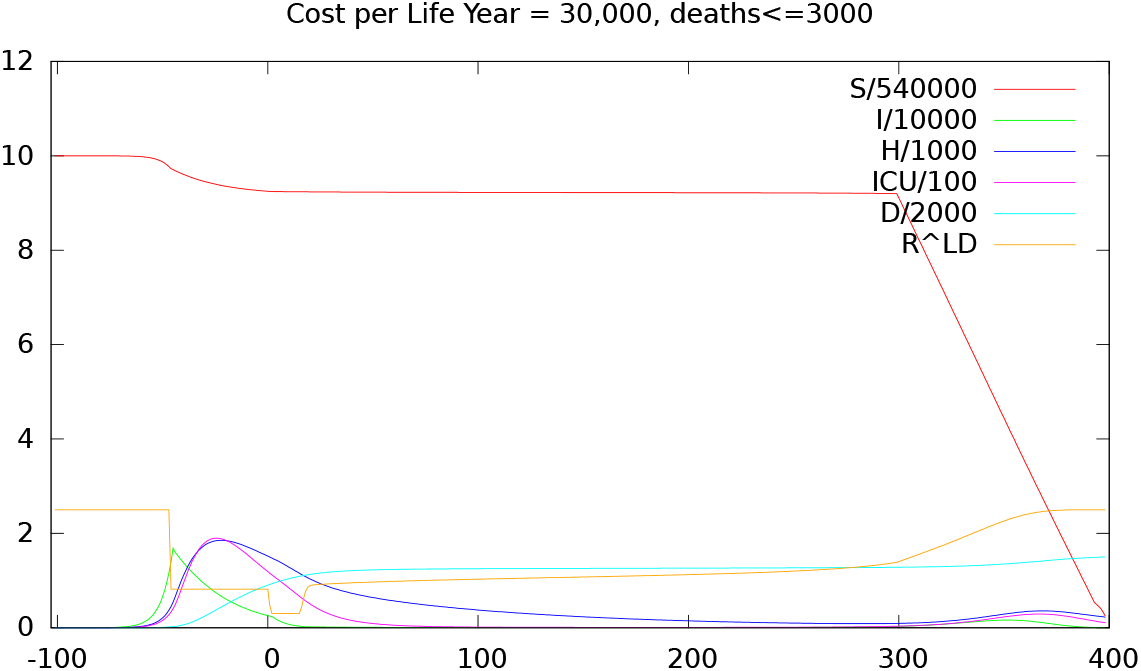

In this case the plot looks identical to the one for QALY=£300,000 and indeed mathematically these two formulations are equivalent. We note that this plot corresponds to a cost of £37.1bn with the only difference to the earlier case that the loss of 3000 lives is now valued at £0.63bn (rather than £6.3bn); the economic damage due to the lockdown is estimated as £36.4bn in both cases.

It should be noted that these final cases corresponds to a strategy of keeping the reproduction rate ℛ close to 1 and completely preventing a second wave until a vaccine is found, which seems to be the strategy favoured at the moment by most western governments.

In summary these policy optimization runs without tracing paint a rather gloomy picture: there seems to be a choice between only three options: to release lockdown regardless of overflowing hospitals, to release lockdown after it is made sure that hospital capacities are not overloaded, both resulting in an unacceptably high death toll, or to keep the lockdown in place until a vaccine is found at an unacceptable economic cost.

### 5.1. The Effect of Testing and Tracing

We now give the optimization the option to reduce ℛ by testing and tracing combined with a degree of lockdown. As outlined earlier our default assumptions are that we can never find more than 50% of the currently infectious people to trace, tracing is only 90% effective and we can remove a maximum of 900 people a day from the infectious pool by tracing. To test one infectious person and trace and isolate their contacts costs £6500. We also use the default QALY value of £30,000 and impose no limit on lives lost. The optimal intervention policy and resulting disease dynamics are displayed below.

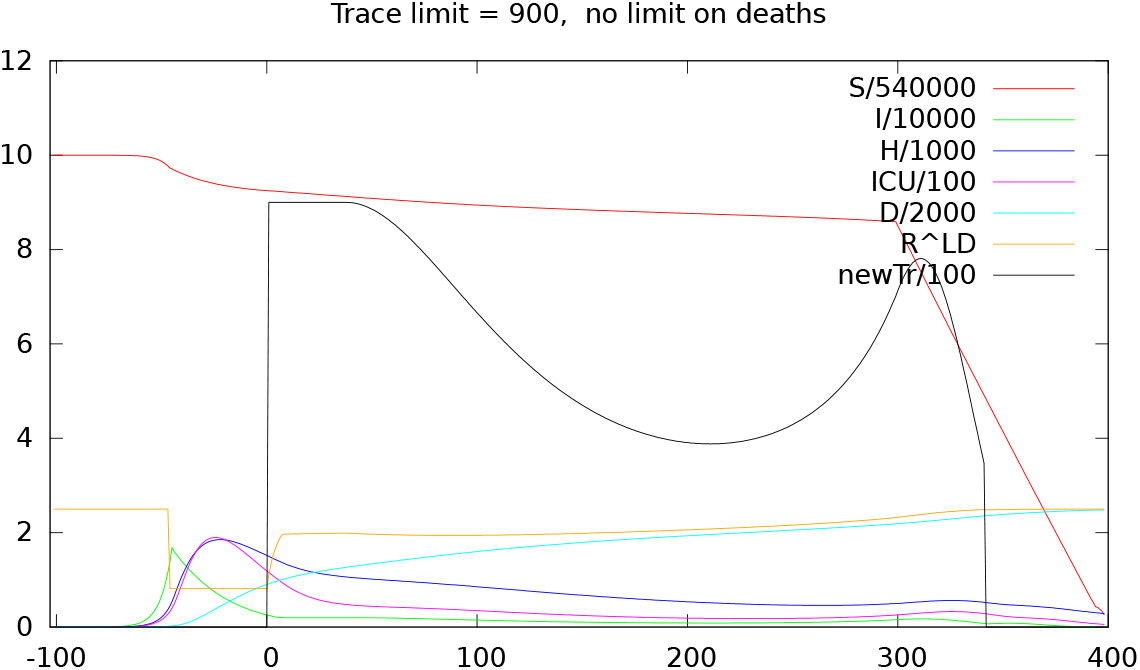

The quantities on the plot are as before but now additionally show newTr, the number of infectious people who have their infectiousness reduced by tracing per day. This policy allows the lockdown levels to be relaxed to about ℛ^*LD*^ = 2.0 (which may be achievable by social distancing measures[4]) almost immediately, while keeping total deaths below 5, 000. The total cost of this scenario is £6.241bn (3.6% of GDP), thus cheaper than any of the others. Tracing is used to its full capacity until about *t* = 50 at which point it falls because the number of infectious people falls below 1800 and thus tracing is limited by the 50% limit. Tracing increases again just before the start of the vaccination programme in line with the final relaxation of the lockdown.

Of course the death toll of 5,000 exceeds the limit of 3,000 that we have imposed in the last section, so in order to compare results we re-run the tracing model but with a death toll limit of 3,000. This scenario has a similar intervention shape as the one before. Lockdown measure are not relaxed as far as before (now to around ℛ^*LD*^ = 1.8) which together with tracing quickly reduces the number of infectious people to below 1000 at *t* = 26. Due to the very low infection levels, few tracers are needed to keep the disease under control. As before lockdown is relaxed and tracing increased again just before the start of the vaccination programme. The cost of this scenario is £7.223bn of which £6.262bn is due to the economic damage, £0.62bn due to lives lost and a mere £0.34bn due to tracing. This should compared to the cost of £37.1bn needed to achieve the same limit on death toll without tracing, a factor of 5 improvement. Note that this decrease of the total cost when using tracing is in line with the more theoretical analysis in Section 2.

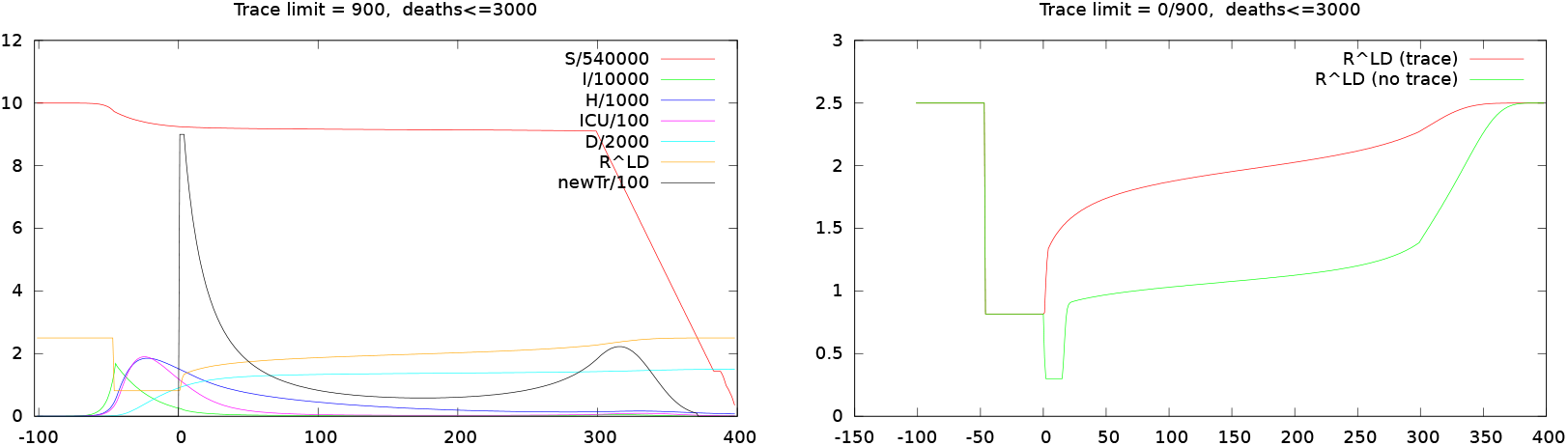

We also compare the optimal 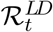 that would need to be achieved by lockdown measures for the cases with and without tracing but an imposed limit of deaths of 3,000. The difference is striking. We can observe that in both cases the lockdown measures are gradually relaxed during the middle of the intervention period (*t* ∈ [50, 250]). We think this is due to the underlying exponential nature of the disease spread: any infection prevented at an early time will prevent a large number of infections in the future, this effect is more pronounced the earlier the initial infection is prevented; hence there is a stronger reason for strong control at the beginning rather than later.

Note that in the tracing case, the lockdown measures are relaxed to about 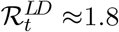 (which is reached exactly at *t* = 70, almost the same day that 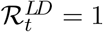 for the case without tracing). This ℛ^*LD*^ = 1.8 is exactly that predicted by the plot in Figure 1 for 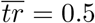 and *η* = 0.1.

**Figure 1.**
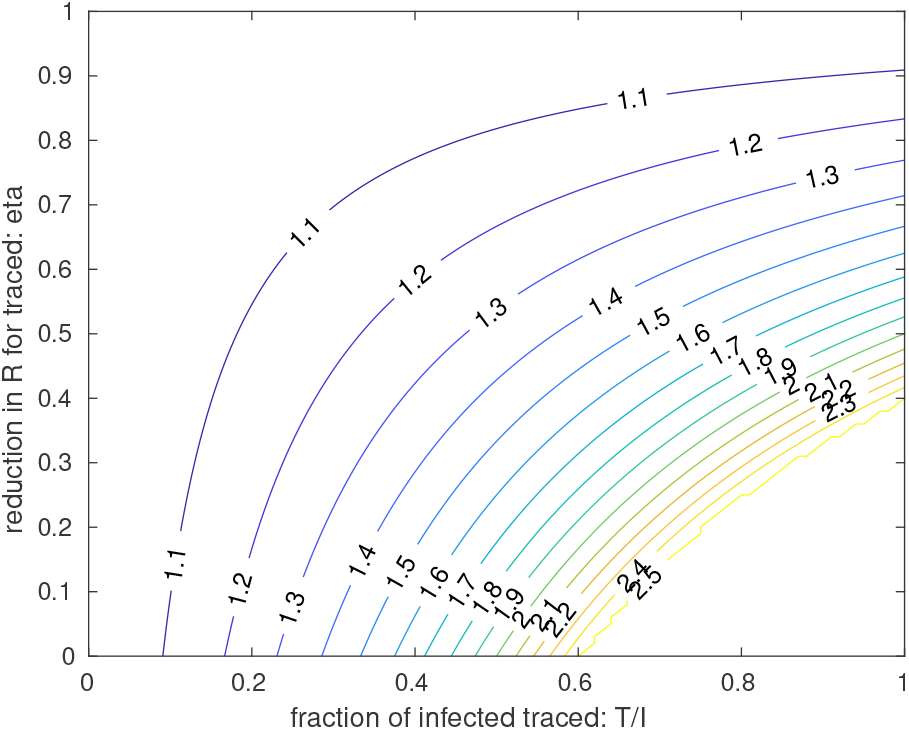
ℛ^*LD*^ needed to keep the observed reproduction number ℛ = 1 for various choices of tracing parameters *T*/*I* and *η*. Plot assumes ℛ0 = 2:5 and *s* = 1.

### 5.2. Robustness of Results

While the results presented in the last section clearly show the advantages of using tracing, we have made quite a few assumptions in obtaining them and one natural question is how robust the results are regarding changes in parameters.

Firstly, one could ask inhowfar the optimal intervention strategy is biased by the model knowing that a vaccination programme is available at time *t* = 300. In order to explore this we have delayed the vaccination programme by another year (now starting at day *t* = 600). We also proportionally increase the limit on the death toll to 4000 (in the previous runs about 1000 deaths were allowable for *t* ∈ [1, 300], we allow another 1000 for *t* ∈ [301, 600]). This results in a very similar optimal policy as before (shown below). The intervention in both lockdown and tracing is slightly more pronounced at the beginning than before.

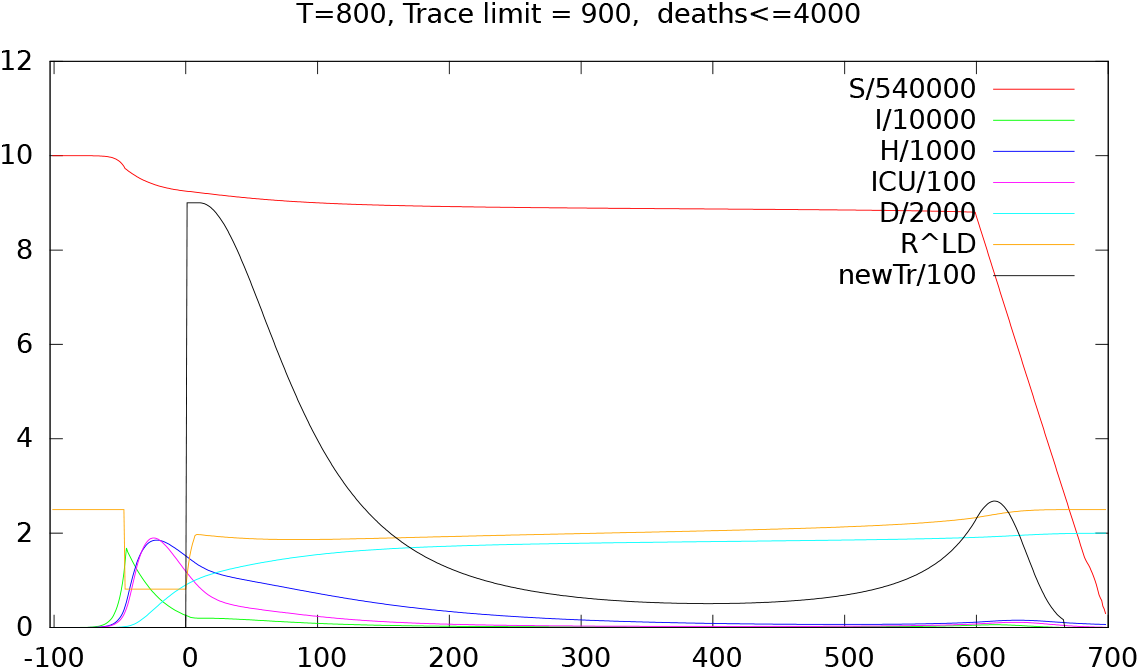

As the next test we have investigated the effect that the required deaths limit has on the possible relaxation of the lockdown measures and the tracing needed. The results are in the two figures below.

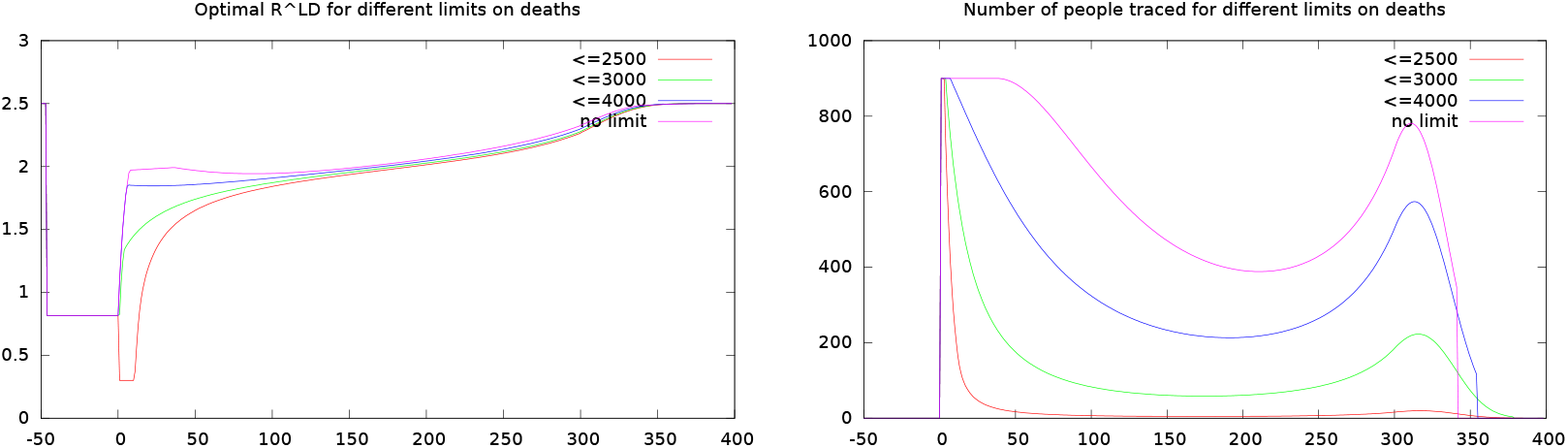

From the first plot we see that the final death limit has almost no effect on the optimal ℛ^*LD*^ value over most of the planning interval (*t* ∈ [100, 300]), this is in line with our earlier analysis that the allowable ℛ^*LD*^ only depends on the parameters of the tracing. There is however a difference in how fast the lockdown is relaxed at time *t* ∈ [0, 100] depending on the death limit. If there is no limit the lockdown is relaxed immediately. If there is a very tight limit, however, the lockdown is initially even tightened for about a week until the number of infectious people is sufficiently decreased and then relaxed.

The plot on the right shows the number of people traced over time for different deaths limits. Somewhat counter-intuitively the number of people being traced is lower (significantly so) when the deaths limit is lower. The reason is that for a tight deaths limit, there is initially a severe lockdown, so that only very few infectious people remain. Subsequently only very few people can be traced. We see that in all cases the number of traced people, in line with the number of infectious people, increases when the lockdown is relaxed in anticipation of the start of the vaccination programme at *t* = 300.

We will now investigate the sensitivity of the results with respect to changes in the tracing effectiveness (1 − *η*) and the fraction of infectious people traced 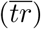. We first limit the fraction of traceable people at 50%, impose a deaths limit of 3000 and vary *η*. The plot on the left shows the optimal intervention strategy in terms of ℛ^*LD*^, whereas the plot on the right shows how the associated costs (in £1m) change as *η* changes.

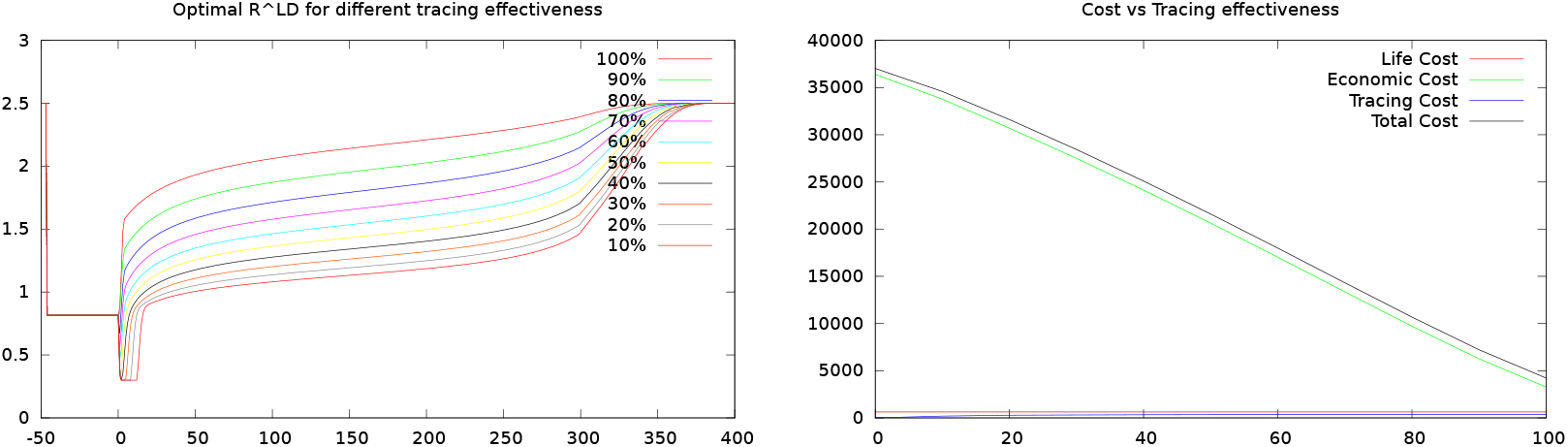

We see that in line with the earlier analysis a higher value of tracing effectiveness results in a larger ℛ^*LD*^ (less severe lockdown measures). We also see that there is a steep decline in associated costs as the tracing effectiveness is increased.

The next plot keeps the tracing effectiveness at 90% (*η* = 0.1) and varies the fraction of the infectious people that can be traced. Again the results are in line with the earlier analysis. Note that with *η* = 0.1, for 70% and 80% of infectious traced, tracing alone can contain the spread of the disease without any other measures. Subsequently in both these cases the lockdown is relaxed almost immediately to ℛ^*LD*^ = 2.5. Again costs decrease rapidly with higher proportion traced, until they flatten out when tracing alone would be sufficient to control the disease.

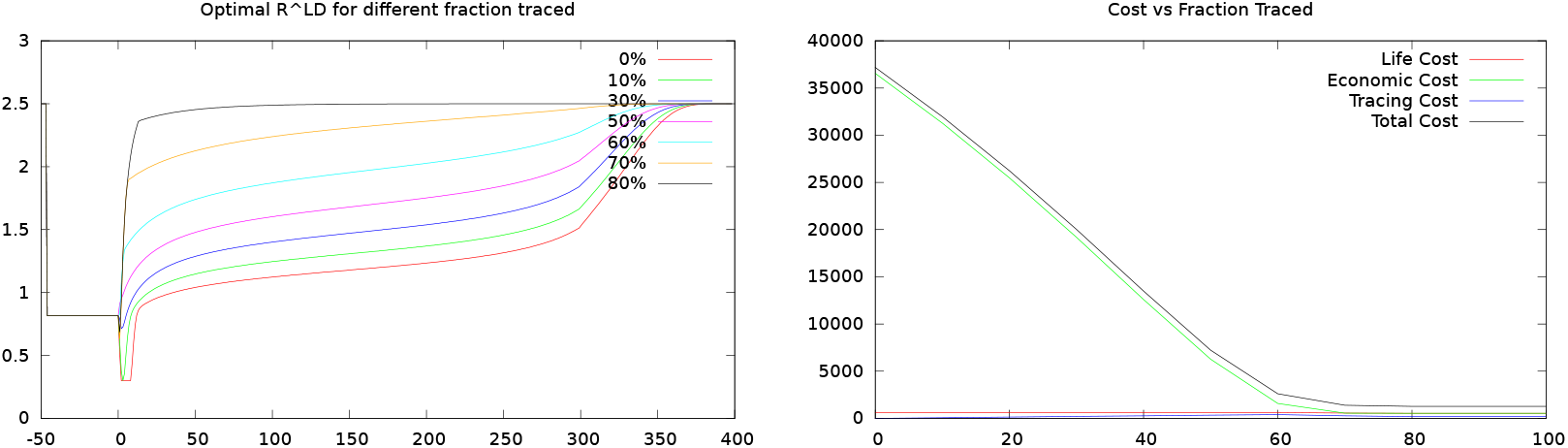

In everything so far we have assumed that there is the capacity to trace and follow up contacts of up to 900 identified infectious people every day (in Scotland alone). As Scotland was testing about 2500 people a day in mid May, this value of traces may be seen as overly optimistic. In the final plot we therefore investigate the effect of setting the capacity of number of people whose contacts are traced per day to different values.

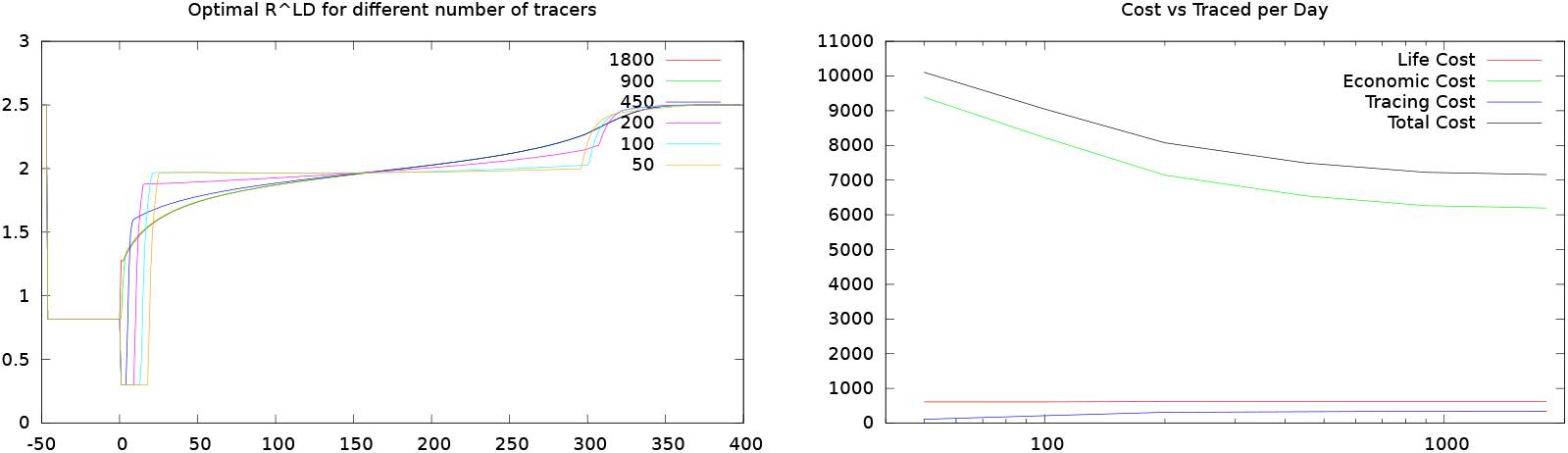

Surprisingly the optimal ℛ^*LD*^ does not depend on the tracing capacity, and even for a capacity of only 50 people’s contacts traced per day we can still relax the lockdown to ℛ^*LD*^ = 1.8 (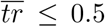, *η* = 0.1). What does change is that for large tracing capacity the lockdown can be relaxed immediately, whereas for small capacity the lockdown is initially tightened for up to 20 days. This is again in order to get the number of infectious people so small that even the small available number of tracers can deal with the cases that may arise once lockdown restrictions are eased. Another effect of a smaller tracing capacity is that the lockdown level stays constant rather than gradually loosening as for the default case.

Note that due to the tighter lockdown needed initially for the case with the small number of tracers, the total cost of the policy increases from about £7bn for 1800, 900 and 450 tracing capacity (with only a small increase) to about £10.5bn when the capacity is only 50 people per day.

In order to have a fuller picture we give the complete analysis of the optimal policies and disease dynamics for the two extreme cases of 1800 and 50 per day for the tracing capacity (note that the plot on the right has a different scale). We see that when 1800 people can be traced, these are not actually used. For only 1200 people contacts are traced at the beginning (since there any only about 2400 infectious people) and after that the number of people traced falls quickly together with the number of infectious people. If tracing capacity is only 50 people per day, then this is fully used for the full period (indeed until after the start of the vaccination programme).

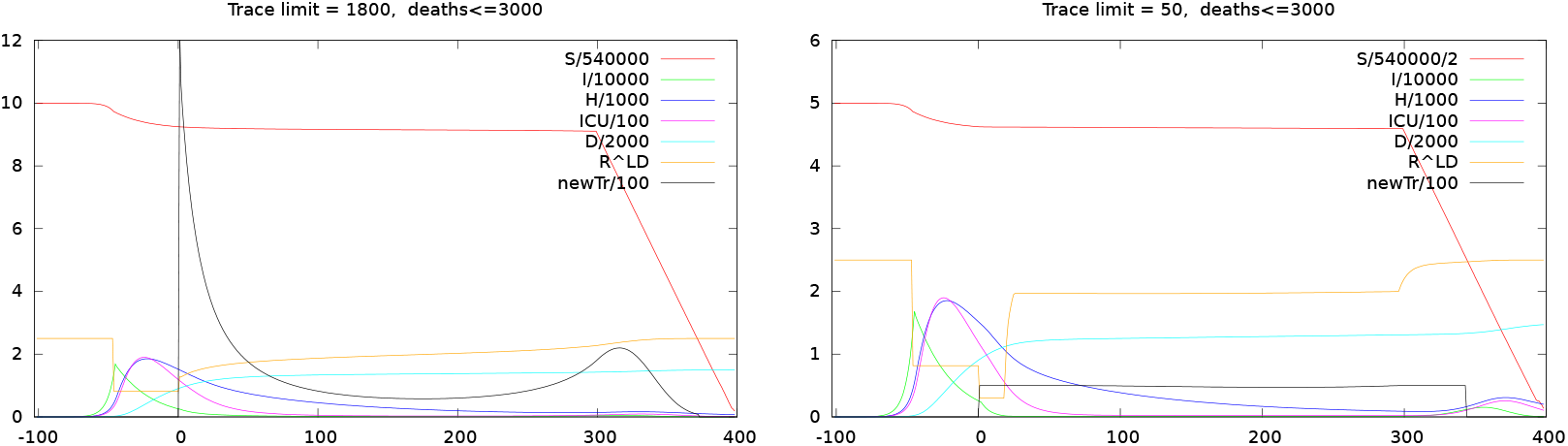

We should point out that the results for the smallest tracing capacity (50) are likely overly optimistic. They rely on the fact that the policy optimization makes optimal decisions and is thus able to predictably keep the number of infectious people at manageable levels throughout. In practice this will not be possible, partly because especially if numbers are low, the exact number of infectious people will not be known, and secondly it would be expected that there may be occasionally sudden outbreaks of many new cases (so-called superspreader events) that prevent keeping the number of infectious at very low constant levels.

In summary we see that tracing has an important role to play for any assumed level of tracing effectiveness, fraction of infectious people traced and daily tracing capacity. There is a marked reduction in overall intervention costs if the tracing effectiveness is high and a larger number of the infectious can be identified and their contacts traced. In principle these results also hold if the absolute number of traces that can be performed per day is low, although in that case the total number of infectious people must have been reduced sufficiently by lockdown measures for tracing to be effective.

## 6. Conclusions And Outlook

We have presented a policy optimization model that can be used to dynamically determine the optimal intervention strategy to control the spread of the COVID-19 epidemic over time. In addition to lockdown measures, the model can also decide to use testing and tracing. We have further given an analysis that shows testing and tracing in a combined strategy can be many times more cost effective at reducing the ℛ number than lockdown measures alone.

When running the model with parameters originating from a least squares fit to the available data for Scotland, we find that testing and tracing results in strategies with a lower death toll and lower costs than without tracing. For high values of tracing effectiveness and fraction of infectious people traced the differences compared to an intervention strategy without tracing are striking. In the cases where the theoretical analysis suggests that tracing alone would be able to contain (or nearly contain) the spread of the virus, the policy optimization confirms that no (or little) additional lockdown measures are needed. However, even for low tracing effectiveness, the model shows a marked reduction in lockdown measures and associated economic costs compared to a model without tracing. This is in contradiction to the popular claim that tracing would need to find at least 60% of infectious people in order to be worthwhile.

We should note that there are many assumptions and simplifications in our model and despite our best efforts the data that we used and inferred through the fitting process has large error bars associated with it. The model therefore cannot be used to make any quantitative predictions about the future evolution of the disease or about the optimal intervention strategy. We also want to point out that the model is able to make optimal decisions, and consequently at early times in the intervention cycle will assume that optimal decisions are made later, (*e*.*g*. with regard to severity and timing of any future lockdown interventions). This will result in the predictions from the model to be overly optimistic. Also the model is deterministic in that it assumes full knowledge of the disease parameters (in particular the proportion of the population already exposed to the disease), something that is not known (or will only be known at a later point in time) in practice.

We have, however, done various robustness tests of our results with changed parameters and are confident of the main qualitative result: mainly that effective tracing is a crucial part of any successful intervention strategy to control the epidemic will hold true in the real life setting.

An obvious extension to our study would look at how robust the results are with respect to uncertainty regarding disease parameters, and inexactness of implementing the planned policies in the future.

## Data Availability

All data is publicly available and sources have been referenced in the manuscript.

## Footnotes

1 There is an unfortunate clash of notation here. ℛ_0_ in this context is the basic reproduction number, not the number of recovered people at time t = 0. Since the use of the letter R for both concepts is widespread we dierentiate them by font: ℛ_0_ is the basic reproduction number and *R*_*t*_ the number of recovered people at time *t*.

2 Date of fitting the data: 11 May 2020

3 11 May 2020

## Notes

### Competing Interest Statement

The authors have declared no competing interest.

### Funding Statement

No external funding.

